# Improving Fabric Face Masks: Impact of Design Features on the Protection Offered by Fabric Face Masks

**DOI:** 10.1101/2021.01.21.20228569

**Authors:** Eugenia O’Kelly, Anmol Arora, Sophia Pirog, James Ward, P John Clarkson

## Abstract

**Objective:** With much of the public around the world depending on fabric face masks to protect themselves and others, it is essential to understand how the protective ability of fabric masks can be enhanced. This study evaluated the protection offered by eighteen fabric masks designs. In addition, it assessed the benefit of including three design features: insert filters, surgical mask underlayers, and nose wires.

**Methods:** Quantitative fit tests were conducted on different masks and with some additional design features. An array of fabric masks were tested on a single participant to account for variability in face shapes. The effects of insert filters, surgical mask underlayers and nose wires were also assessed.

**Results:** As expected, the fabric masks offered low degrees of protection; however, alterations in design showed significant increase in their protective ability. The most effective designs were multi-layered masks that fit tightly to the face and lacked dead space between the user and mask. Also, low air-resistance insert filters and surgical mask underlays provided the greatest increase in protection.

**Conclusions:** Our findings indicate substantial heterogeneity in the protection offered by various fabric face masks. We also note some design features which may enhance the protection these masks offer.

## Introduction

The COVID-19 pandemic has inflicted unprecedented strains on global supply chains of all personal protective equipment (PPE), particularly N95 and FFP3 masks. Due to these shortages, there have been coordinated government actions aimed to discourage the use of hospital-grade masks by members of the public in order to reserve their use for clinical application. The general public, as well as clinicians in circumstances when there is a deficit of suitable PPE for clinical use, are encouraged to use fabric face coverings to help limit the spread of the SARS-CoV-2 virus. ^*1,2*^ Unfortunately, the body of research assessing the optimal design and construction of fabric face coverings is limited, with most research studying the filtration of fabrics and other potential materials rather than design features.^*3,4,5*^ Many who rely on fabric masks understandably wish to optimize the protection their fabric face mask offers, not only for others but also for themselves.

Multiple methods to improve fabric mask protection are being widely used and marketed. Fabric masks are often made with a pocket into which a replaceable filter can be inserted. High-filtration materials such as particulate matter (PM2.5) or HEPA (High Efficiency Particulate Airfilters) are used in an attempt to increase the protection fabric masks offer; however, given the poor fit of many fabric masks, it is unknown how much benefit such filters impart. A similar approach involves wearing a fabric mask over a surgical mask. Fabric masks often include flexible nose wires in the hope of improving fit. While these design elements have come into common use, there has been little analysis of their impact on a mask’s protective ability.

A mask’s protective ability, as defined by its ability to block particles from invading the inside of the mask, is determined mainly by the filtration ability of its materials and the fit of the mask on the wearer.^*6*^ While several studies have investigated the filtration ability of common fabrics^*3,4,7,8,9*^ little work has been done on the fit of fabric masks. While the importance of fit has long been understood to be one of the primary factors in determining the effectiveness of face masks^*10,11*^, limited research on fabric face mask fit has been conducted.

While fabric masks can never offer the same protection as hospital-grade masks, increasing fabric mask protection is beneficial for both the wearer and those in close proximity. For the wearer, limiting the number of particles which penetrate into the inside of the mask decreases the likelihood of harmful particles affecting the wearer, and potentially decreases the inoculum of SARS-CoV-2.^*12*^ For others in close proximity, a mask with better fit and materials will be more likely to trap the wearer’s respiratory emissions.

This current study seeks to assess (a) the protection offered by multiple fabric face masks designs and (b) how design features such as insert filters and nose wires impact protection.

## Materials and Methods

To determine the amount of protection various fabric masks offered, the method of quantitative fit testing was used. Quantitative fit testing is the most rigorous and objective form of mask fit testing commonly available. It is frequently used in industry and healthcare to assess the fit of respirators, including N95 and FFP3 masks. During quantitative fit testing, air samples are continuously taken from both inside and outside the mask. Particle counts from both air samples are then compared to provide a ‘fit factor’ or ‘protection score’, which represents the proportion of particles the mask has blocked. The concentration of particles outside of the respirator are divided by the concentration of particles inside the respirator via an OSHA protocol.^*12,13*^

When testing masks with less than a 99% efficiency, scores range from 0 to over 200, represented as 200+. According to OSHA guidelines, a properly fit N95 respirator must score above 100. Previous studies have shown basic fabric masks have a fit factor of around two.^*6*^

Quantitative fit testing scores represent both fit and filtration; as, fewer particles inside the mask may be a result of better fit and/or better filtration. In the case of N95 masks, low scores can be safely assumed to be due to a fault in fit only, as the material has been previously tested for its high filtration ability. However, in the case of fabric masks, where both material and fit are variables, the fit factor score represents a measure of protection offered from the mask as a whole - it’s combined fit and filtration. High fit factors indicate greater protection than low fit factors, with fewer particles having penetrated the material and/or entered around gaps in the mask.

Participants followed OSHA protocol 29CFR1910.134 during quantitative fit testing. In accordance with this protocol, participants complete a series of activities and motions designed to mimic the scope of occupational activity. These activities include normal breathing, deep breathing, turning the head, and bending over. Quantitative fit tests were conducted using a Portacount, TSI, Minnesota, model 8038+ capable of evaluating masks with less than 99% efficiency.

The first goal of this study was to evaluate the range of protection various fabric masks can offer. Previous studies have performed quantitative fit testing on basic fabric masks and recorded scores of approximately two^*6*^, but the masks tested in these studies were few in number and basic in design. To assess a wider range of masks, one member of our experimental team tested seventeen masks of various construction methods and materials. During this analysis, all masks were tested on a single participant to control for variations in facial features. Thus scores generated should not be taken to represent the amount of protection another individual should expect from any one mask, but instead interpreted as an estimate of how well one fabric mask functioned in relation to other fabric masks. Some difference in fit should be expected between participants.

Multiple members of the team have observed members of the public wearing fabric masks on top of surgical masks. This might incur two benefits. The surgical mask may act as an additional filtration layer and the fabric mask, if it fits the wearer well, may help compensate for the poor fit of many surgical masks.

To test the benefit of wearing a surgical mask under a fabric mask, a store-bought surgical mask was worn under various fabric masks and quantitative fit testing performed. Two participants were randomly assigned to a fabric mask type during this testing.

Three types of filters were assessed for their potential improvement to the wearer’s safety. These included a PM 2.5 filter, a repurposed paper-type HEPA vacuum filter, and a repurposed ventilation system filter. To test the benefit of different types of filters, the same mask was tried with each type of filter inserted into a filter pocket in the mask. Quantitative tests were performed to assess the degree of added particle filtration each filter created. This portion of the study consisted of sixteen tests on four mask models using three participants randomly assigned to a mask type.

Each filter type offered different breathing resistances and filtration benefits.

The vent filter provided extremely low breathing resistance but was intended only to help collect dust. It was not certified as a particulate air filter. The paper-type HEPA (high efficiency particulate air) vacuum filter was extremely difficult to breathe through but promised HEPA levels of filtration. The PM2.5 filter had a breathing resistance slightly higher than the vent filter and promised high filtration benefits. The purpose of including filters with various degrees of breathing resistance was to observe if high-resistance filters, such as the vacuum HEPA filter, would incur less benefit than might be expected by their filtration ability alone. It would be expected that a mask with high air resistance will encourage increased leakage during breathing.

Many insert filters offered on the market are not large enough to cover the entirety of the mask. Critically, they may cover the mouth area but not cover the nose area. To assess how important the size of the filter was, we tested a filter which covered 100% of the mask area and a filter which covered approximately 50% of the mask area. Each 50% filter was cut to the size of a store bought PM2.5 filter. In some masks, this filter covered slightly more than 50% of the area while in other masks it covered less. A total of nine tests were performed by two participants randomly assigned to a mask type using repurposed vent filters and repurposed paper vacuum bag filters.

To evaluate the fit improvement incurred by the inclusion of a nose wire, five masks were tested with and without nose wires. A 3mm aluminium armature wire was used as nose wire for the home-made masks tested. Two participants of the study team were randomly assigned to test various mask models with and without nose wires.

Limitations to the Study: Experimental testing on filter size may have been influenced by the placement of the quantitative fit testing machine’s mask sample valve through which interior air samples are taken. In all cases, the valve was placed near the middle of the mask breathing area as instructed by fit testing protocol.^*13*^ Depending on the design of the mask, this most often resulted in the valve being (a) closer to the mouth than the nose and (b) centered in the middle of the 50% filter. This placement may have resulted in some artificial benefit for 50% masks.

## Results

### Fabric Mask Fit

The seventeen masks tested included both commercial masks and homemade masks. Photographs of each mask and a description of its design can be found in Figure 1.

**Fig 1:**
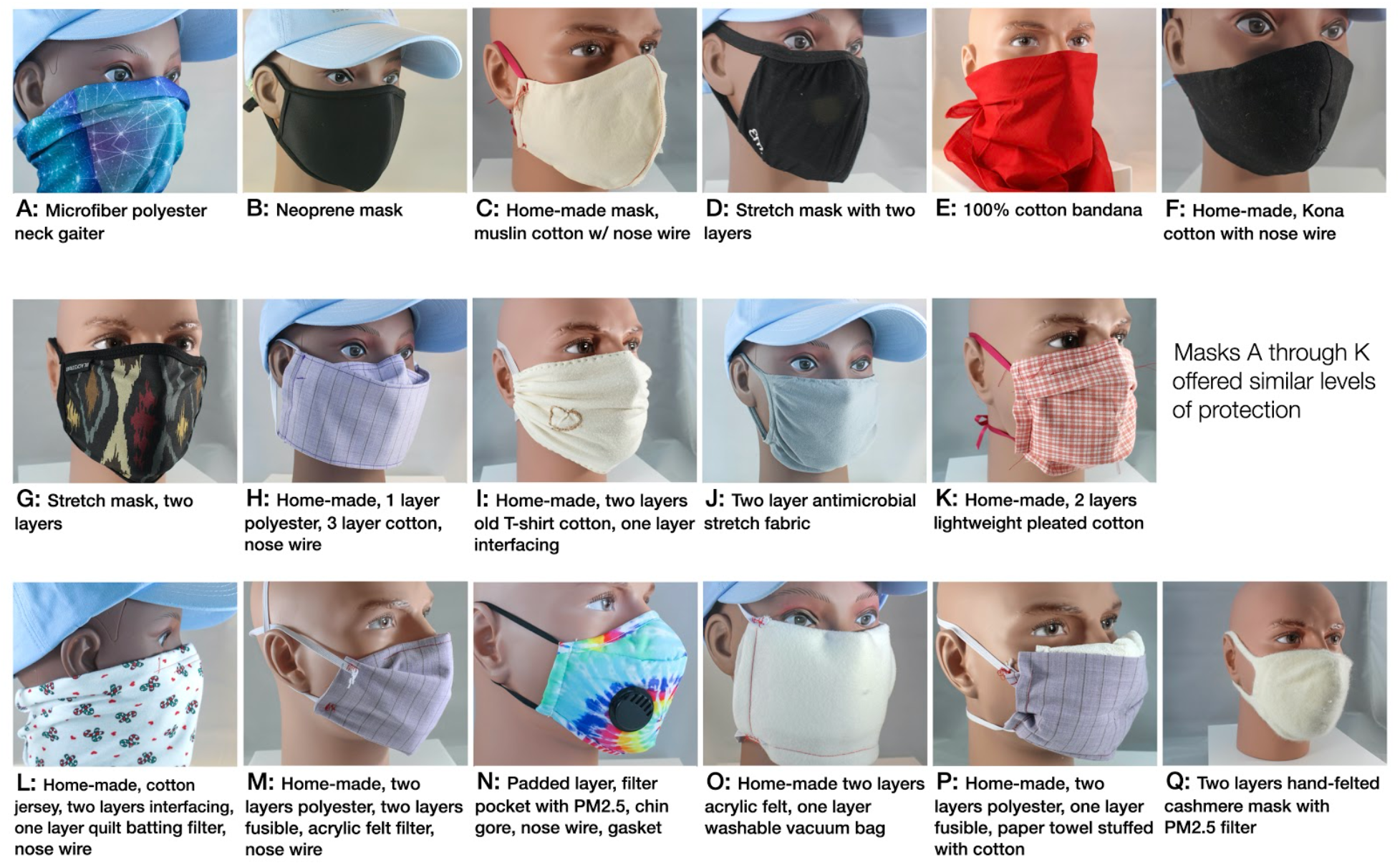
Photographs of the seventeen masks assessed for their protection factor on mannequin heads. Details of the construction and materials of each mask are included whenever possible. Masks that were home-made are identified.

**Fig 2:**
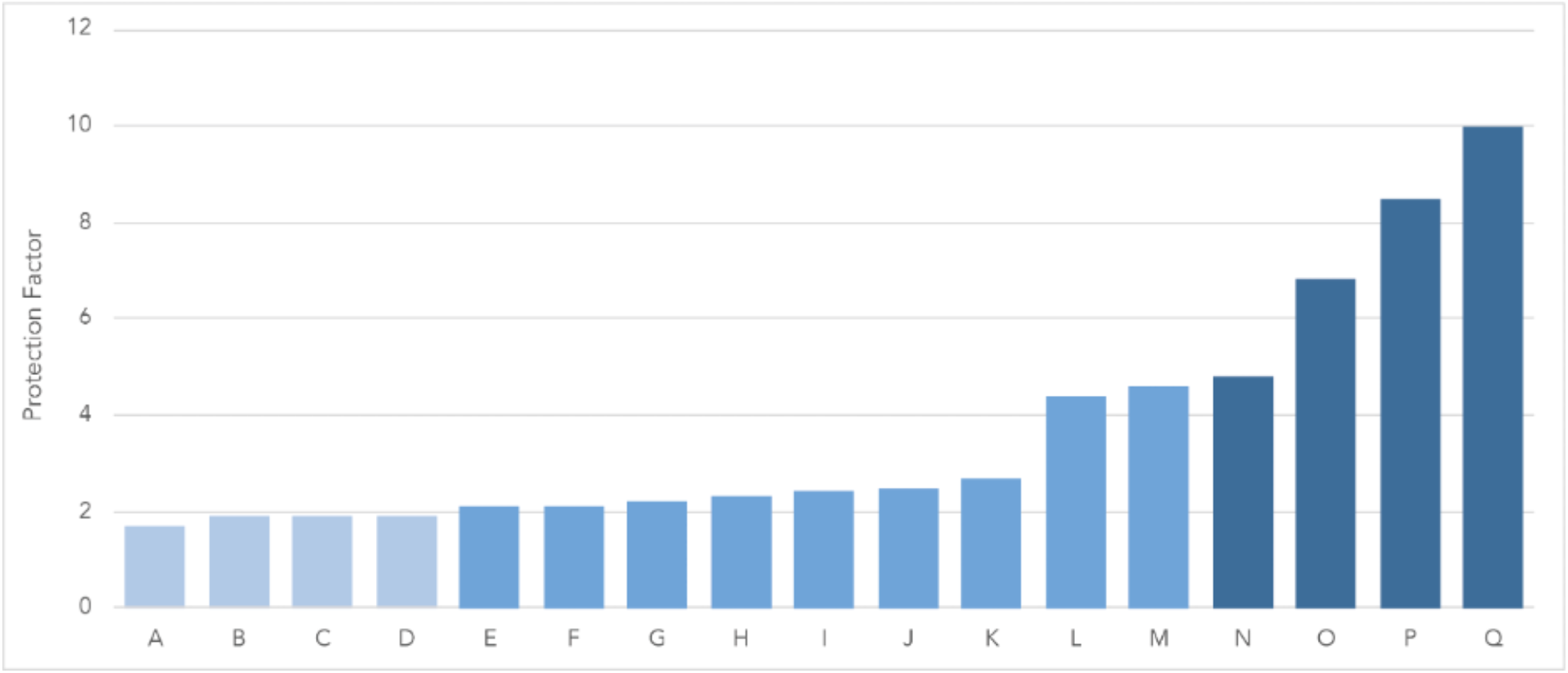
Protection factor of the seventeen tested fabric masks.

The majority of fabric masks (A-K) provided similar, low levels of protection between 1.7 and 2.7. These masks generally consist of one or two layers of fabric with no insert filter. The exception is Mask H, which had a complex construction with multiple layers, but a gaping fit which failed to conform to the sides of the face. The low scores of Mask H indicate that fit may be more important than material filtration when determining the protection a mask offers. With the exception of Mask H, Masks A-K were closely fit to the face and had small amounts of space between the face and mask, usually 1-3cm.

The second group of masks, Masks L, M, and N provided approximately twice the level of protection with fit factors of 4.4, 4.6, and 4.8. Each of these masks included some type of filter, either a home made filter as in the case of Mask L or a commercial such as the PM2.5 filter used in Mask N. These masks had good fit with no noticeable gaps. Masks M and N had approximately 3-4cm of dead space between the face and mask.

Masks O, P, and Q offered increasing levels of protection of 6.8, 8.5, and 10 respectively. Each of these masks were characterized by a very tight fit to the face accompanied by a noticeable increase in breathing resistance. In each of these masks the material was pulled flush against all tissues of the face. There was no dead space between these high-performance masks and the face. Masks O, P, and Q had noticeable thickness, and when tied tightly on this bulk of the mask allowed the mask to contour around the end of the nose.

Mask P was highly effective but difficult to breathe through. One participant described it as being suffocated by a scratchy diaper. Mask Q provided the highest protection with a score of 10. It fit tightly but not uncomfortably to the face and was made of two layers of hand-felted cashmere with a PM2.5 filter inserted between.

### Wearing a Surgical Mask under a Fabric Mask

Wearing a surgical mask under a fabric mask proved to increase protection significantly (see Figure 3). In some cases, protection was doubled, with an average increase of 2.5. Masks which fit more tightly to the wearer’s face provided greater benefits than those which fit poorly. In only one case was no improvement noted. For this test, Mask D was tested with a PM2.5 filter inserted into the pocket. Adding a surgical mask below this mask did not result in any change to protection factor.

**Fig 3:**
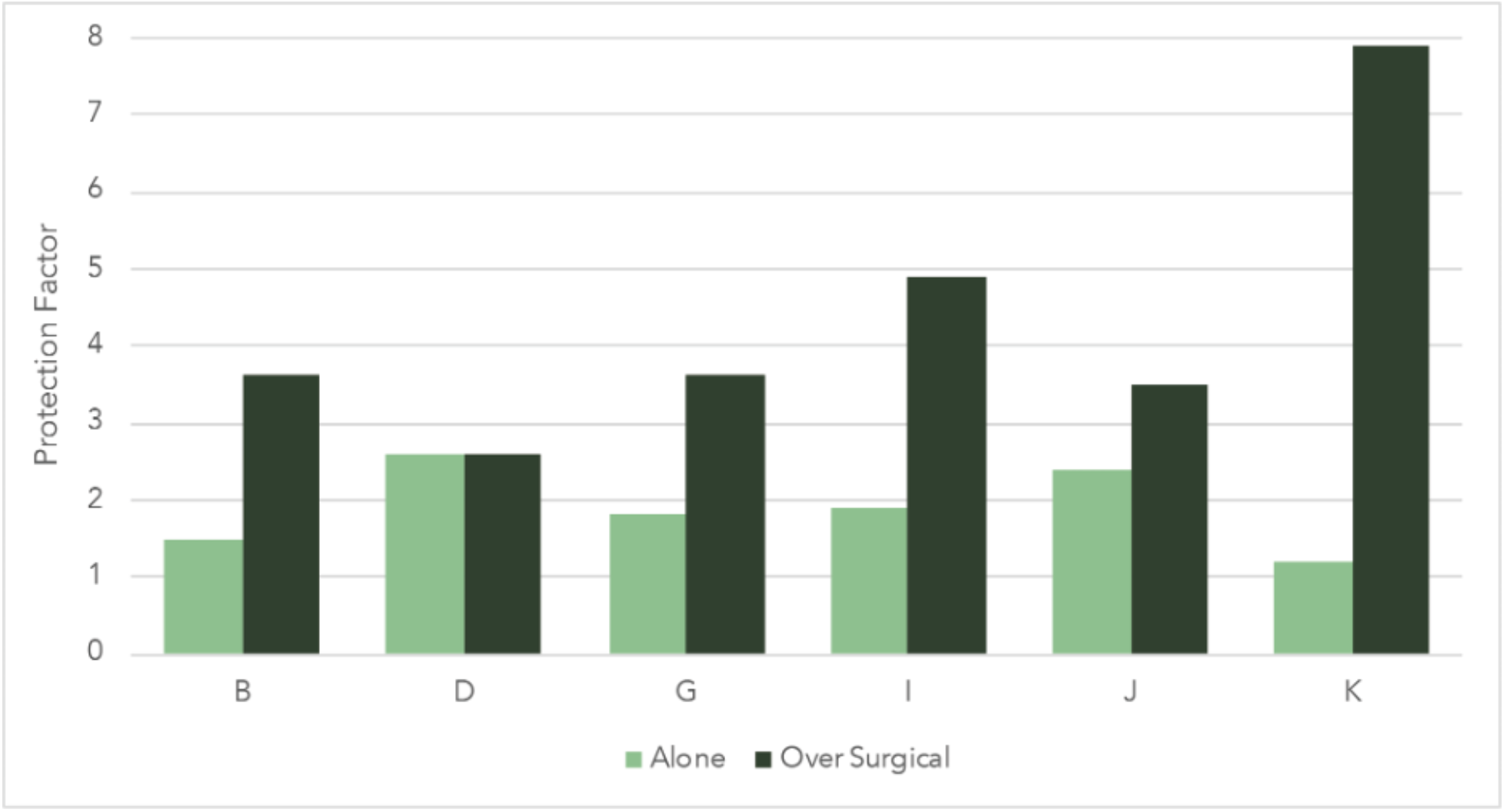
A comparison of fit scores of fabric masks tested alone as compared to when the same fabric mask is worn over a surgical mask.

The greatest improvement came from including a surgical mask under the Mask K, which employed a style similar to a surgical mask, with an improvement of 6.7.

### Size and Type of Filter

Including a filter in a fabric mask was shown to offer clear benefits (See Figure 3). Including a filter half the size of the mask, centered on the breathing area, improved masks by an average of 1.2. By increasing the filter to cover the entire mask, an additional 0.6 score was achieved.

Using a PM2.5 filter improved mask protection by an average of 1.7. The ventilation filter provided lower filtration promises than the PM2.5 or paper-type HEPA filter. However, its extremely low air resistance resulted in a protection increase very similar to that of a PM2.5 filter. The high air-resistance HEPA paper vacuum bag improved masks by an average of only 0.8, despite its high filtration rating. This indicates that low air resistance is critical when choosing a filter for a mask. A filter with a lower filtration ability and low air resistance may be more effective than a high filtration ability and high air resistance.

### Nose Wire

Results from the nose wire tests were inconclusive (figure 5). Not all masks benefited from the inclusion of a nose wire. Participants noted that on some masks, the nose wire could prevent the mask from adapting to movement, thus creating new fit gaps in other areas of the mask. On other masks the nose wire was observed to make a noticable improvement. Our research team observed that nose wires were more helpful on masks with more structure, such as the two panelled KN95 and least helpful on masks with stretch material or those whose material had low rigidity.

**Fig 4:**
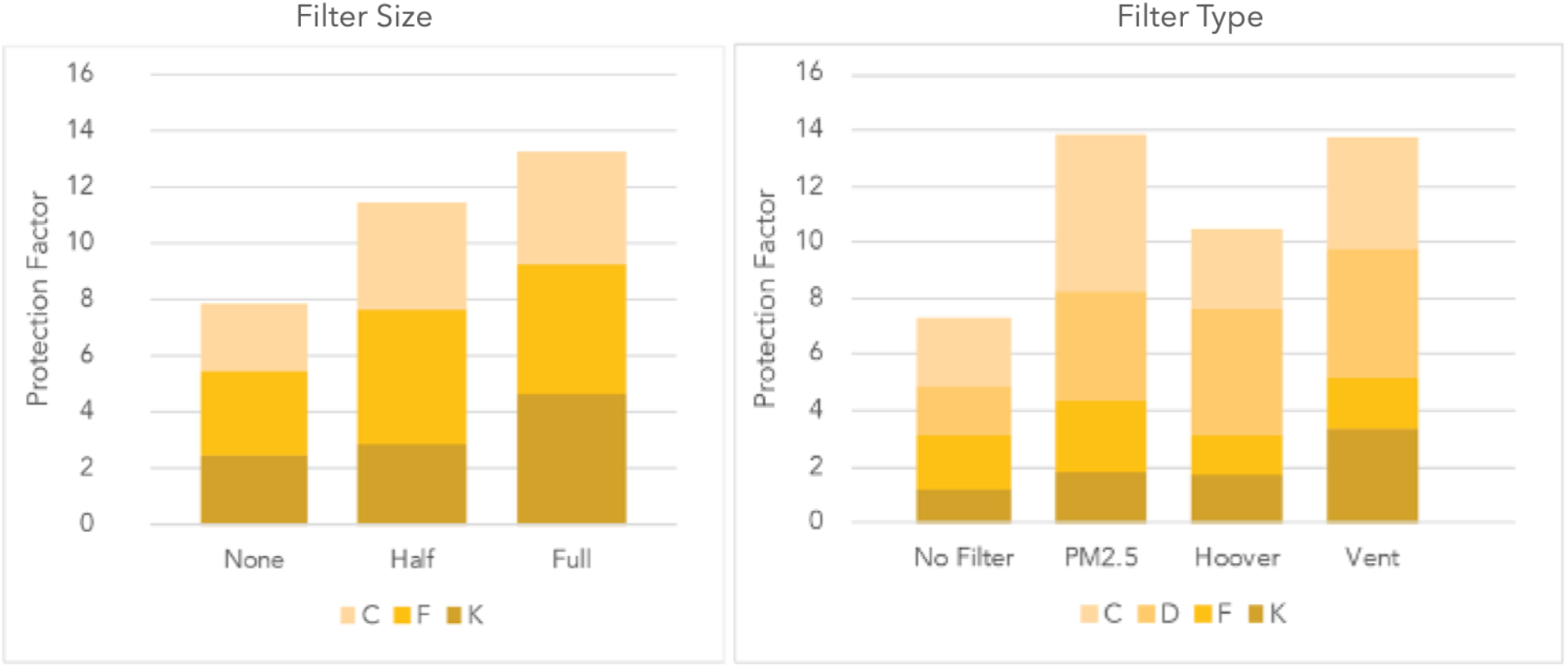
Protection factor improvement by the size and type of filter.

**Fig 5:**
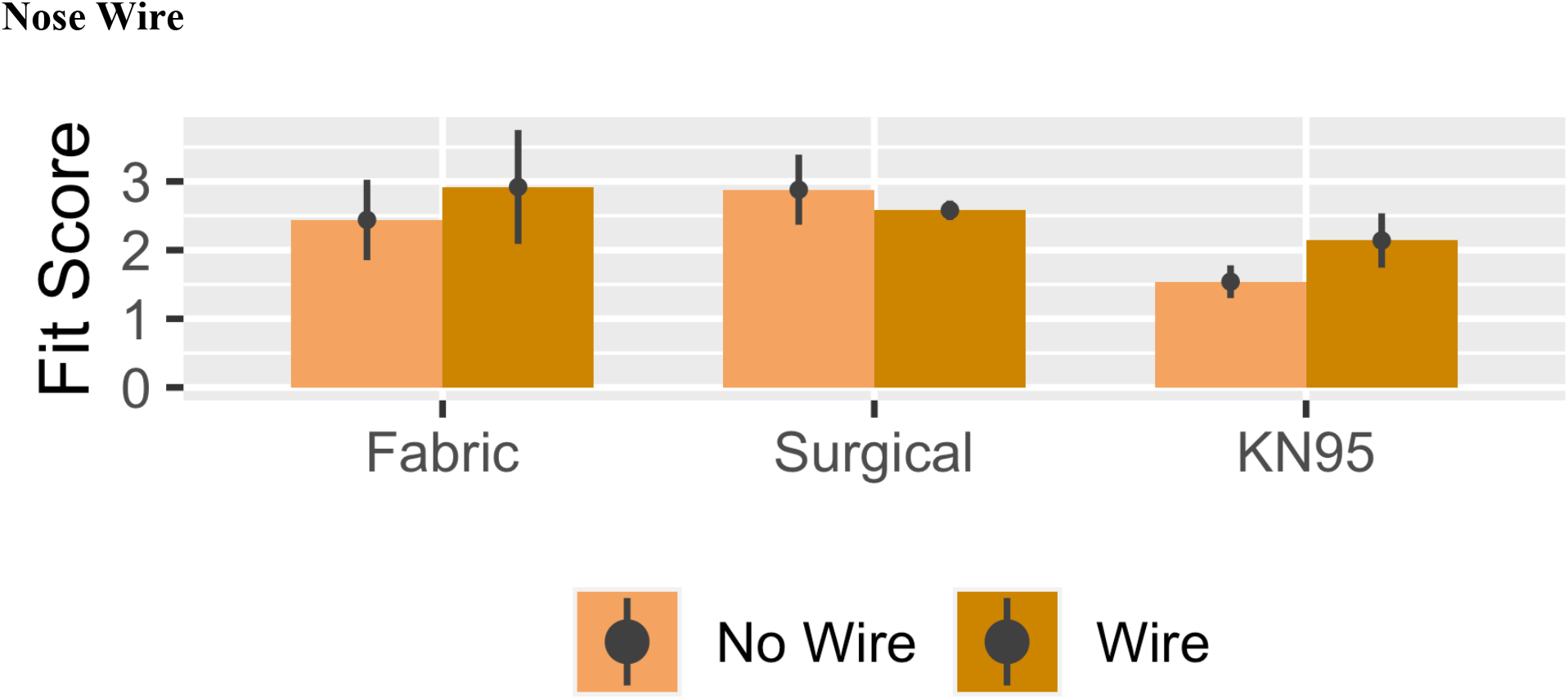
Performance of mask groups with and without wire. Five samples in each group were tested.

## Conclusion

Our findings indicate that the protection offered by fabric masks can range significantly based on fit and construction, with the best mask tested providing five times the amount of protection as low-performing masks. The most protective masks tested were those with multiple layers with a very tight fit. Low air-resistance filters can be inserted into fabric masks to improve protection. Alternatively, a surgical mask can be worn under a tight-fitting fabric mask for significantly improved protection. While fabric masks do not provide the same level of protection as N95 masks, much can, and should, be done to boost the protection they offer.

## Data Availability

Full data from this study is freely availible at the University of Cambridge open repository Apollo: https://doi.org/10.17863/CAM.58287

## Acknowledgements

We would like to thank Corinne O’Kelly for supporting this research. We would also like to thank the many mask designers and home sewers who have shared their patterns, experience, and questions with us.

## Funding

No external sources of funding were used.

## Author contributions

O’Kelly: conceptualization, methodology, writing – original draft preparation, writing – review and editing, project administration. Arora: writing – original draft preparation, writing – review and editing. Pirog: formal analysis, visualization. Ward: conceptualization, supervision. Clarkson: conceptualization, supervision.

## Competing interest

The authors have declared no competing interests.

## Data and materials availably

Data sets are available in the Cambridge University Apollo open access data repository.

## Notes

### Funding Statement

No external funding was received

### Author Declarations

The study has been reviewed by the Cambridge University's Engineering Department Division C ethical review board.

## References and Notes

1. National Centers for Disease Control and Prevention. Use of Cloth Face Coverings to Help Slow the Spread of COVID-19 [Internet]. 2020. Available from: https://www.cdc.gov/coronavirus/2019-ncov/prevent-getting-sick/diy-cloth-face-coverings.html

2. National Centers for Disease Control and Prevention. Strategies for Optimizing the Supply of Facemasks [Internet]. 2010. Available from: https://www.cdc.gov/coronavirus/2019-ncov/hcp/ppe-strategy/face-masks.html

3. Fischer EP, Fischer MC, Grass D, Henrion I, Warren WS, Westman E. Low-cost measurement of face mask efficacy for filtering expelled droplets during speech. Sci Adv. 2020;6(36):2–7.

4. van der Sande M, Teunis P, Sabel R. Professional and Home-Made Face Masks Reduce Exposure to Respiratory iInfections among the general population. PLoS One. 2008;3(7):3–8.

5. O’Kelly E, Arora A, Pearson C, Ward J, Clarkson PJ. Performing Qualitative Mask Fit Testing without a Commercial Kit: Fit Testing Which can be Performed at Home and at Work. Disaster Medicine and Public Health Preparedness. (2020). https://doi.org/10.1017/dmp.2020.352

6. Comittee on Personal Protective Equiptment for Healthcare Personnel to Prevent Transmission of Pandemic Influenza and Other Viral Respiratory Infections: Current Research Issues. Reventing Transmission of Pandemic Influenza and Other Viral Respiratory Diseases - 2010 Update. Washington, D.C.; 2010.

7. O’Kelly E, Pirog S, Ward J, Clarkson PJ. Ability of fabric face mask materials to filter ultrafine particles at coughing velocity. BMJ Open. 2020;10(9):e039424.

8. Davies A, Thompson KA, Giri K, Kafatos G, Walker J, Bennett A. Testing the efficacy of homemade masks: would they protect in an influenza pandemic? Disaster Med Public Health Prep. 2013;7(4):413–8.

9. Rengasamy S, Eimer B, Shaffer RE. Simple respiratory protection - Evaluation of the filtration performance of cloth masks and common fabric materials against 20-1000 nm size particles. Ann Occup Hyg. 2010;54(7):789–98.

10. Huff RD, Horwitz P, Klash SJ. Personnel protection during aerosol ventilation studies using radioactive technetium (Tc99m). Am Ind Hyg Assoc J. 1994;55(12):1144–8.

11. Cooper D, Hinds WC, Price JM, Weker R, Yee HS. Common Materials for Emergency Respiratory Protection: Leakage Tests with a Manikin. Am Ind Hyg Assoc J. 1983;44(10):720–6.

12. Gandhi M, Beyrer C, Goosby E. Masks Do More Than Protect Others During COVID-19: Reducing the Inoculum of SARS-CoV-2 to Protect the Wearer. J Gen Intern Med. 2020;17–20.

13. Occupational Safety and Health Standards. Standard number: 1910.134 - Respiratory Protection. Available from: https://www.osha.gov/laws-regs/regulations/standardnumber/1910/1910.134

